# Long-Term Efficacy and Allograft Loss after Immune Checkpoint Inhibitor Therapy in Solid Organ Transplant Recipients with Advanced Cutaneous Squamous Cell Carcinoma

**DOI:** 10.64898/2026.09.17.26363341

**Authors:** Hannah Wu, Molly Murphy, Corentin Becot, Angela Fragano, Ethan H.S. Lin, Sruthi Surapaneni, Daniel Shirvani, Christine Cimoch, Ross Merkin, Sonia Cohen, Winfred Williams, Kevin Emerick, Howard L. Kaufman, Sophia Z. Shalhout

## Abstract

**Background:** Immune checkpoint inhibitors (ICIs) can provide effective treatment for advanced cutaneous squamous cell carcinoma (CSCC) in solid organ transplant recipients (SOTRs), but long-term oncologic and allograft outcomes remain poorly characterized.

**Objective:** To evaluate long-term efficacy and allograft outcomes after ICI therapy in SOTRs with advanced CSCC.

**Methods:** We conducted a retrospective cohort study of SOTRs with advanced CSCC treated with systemic ICI at Mass General Brigham from 2016-2025. Tumor response, survival, immune-related adverse events, immunosuppression modifications, rejection rate, and graft loss were assessed. Kidney graft failure was analyzed using competing-risk methods with death as a competing event.

**Results:** Among 36 SOTRs, objective response rate was 44.4% (95% CI, 27.9%-61.9%), including 9 complete responses. At 3 years, progression-free, disease-specific, and overall survival were 18.6%, 65.8%, and 32.5%, respectively. Median follow-up was 42.1 months. Five patients experienced graft loss, all kidney recipients; 4 had biopsy-confirmed rejection. The 3-year cumulative incidence of kidney graft failure was 16.7% (95% CI, 2.9%-30.5%).

**Limitations:** Retrospective design, modest sample size, and heterogeneous immunosuppression limited risk-factor analyses.

**Conclusion:** ICI provided long-term CSCC control with a low rate of allograft loss, highlighting the need for longitudinal multidisciplinary management balancing oncologic benefit, graft preservation, and competing mortality risk.

**Capsule:**

- Long-term outcomes remain poorly defined despite responses to immune checkpoint inhibitors in solid organ transplant recipients with advanced cutaneous squamous cell carcinoma.
- A 44.4% objective response rate, 65.8% 3-year disease-specific survival, and 16.7% 3-year kidney graft failure incidence support shared decision-making and coordinated cancer and transplant monitoring.

## Introduction

Solid organ transplant recipients (SOTRs) bear a disproportionate burden of cutaneous squamous cell carcinoma (CSCC), driven in part by chronic immunosuppression and impaired immune surveillance.^1^ The magnitude of this risk is substantial, with CSCC occurring ∼65-to 250-fold more frequently than in the general population.^2–4^CSCC arising in SOTRs is also more clinically aggressive, with increased risks of recurrence, metastasis, and disease-related mortality compared with immunocompetent populations.^4,5^ The use of immune checkpoint inhibition (ICI) in SOTRs presents a distinct therapeutic challenge because restoration of antitumor immunity may disrupt allograft tolerance and simultaneously promote rejection or graft loss.^6–8^

Programmed cell death protein 1 (PD-1)/ligand (PD-L1) blockade with cemiplimab, pembrolizumab, and cosibelimab has become an established systemic therapy for locally advanced or metastatic CSCC, producing clinically meaningful and durable responses.^9,10^ However, SOTRs were largely excluded from pivotal trials that established the efficacy of these agents because of concerns regarding immune-mediated allograft rejection. Consequently, the long-term efficacy and long-term safety of immune checkpoint inhibitors (ICIs) in this population remain less well characterized than in immunocompetent patients.

Clinical evidence supporting ICI use in SOTRs with advanced CSCC remains limited. A systematic review pooled ICI-treated SOTRs with multiple metastatic malignancies across 36 case-report based publications. Only 4 patients had advanced CSCC and 2 experienced graft failure.^8^ In a multicenter retrospective study of kidney transplant recipients treated with ICIs across 7 cancer types, 24 patients had CSCC. The objective response rate in this subgroup was 36.4%, but CSCC-specific graft loss was not reported, and median follow-up was only 14.2 months.^11^ A prospective Phase I study standardized immunosuppression in 11 evaluable kidney transplant recipients with advanced CSCC by cross-tapering to an mTOR inhibitor and administering pulsed high-dose prednisone. Although no graft loss occurred and 46% of the patients achieved an objective response, median follow-up was only 6.8 months.^12^

These reports have made clinical management challenging as evidence of outcomes in SOTR patients with CSCC is largely derived from small cohorts, short follow-up, heterogeneous malignancies, and studies focused primarily on acute rejection rather than graft failure and the longitudinal consequences of treatment. Thus, we sought to evaluate outcomes of SOTRs with advanced CSCC to define the longer-term tradeoff between durable cancer control and preservation of the transplanted organ. By integrating oncologic response and survival with irreversible graft failure, and real-world immunosuppression management, we sought to characterize clinical benefit beyond the initial decision to administer ICI. Further, data from a large tertiary referral center is expected to inform multidisciplinary care for patients who achieve meaningful cancer control but remain vulnerable to transplant-related morbidity and competing mortality. A better understanding of the long-term outcomes of SOTRs treated with ICI may also inform future transplant eligibility for selected patients who achieve durable oncologic remission in the event of irreversible allograft loss.

## Materials and Methods

### Study design and population

We conducted a retrospective cohort study approved by the Mass General Brigham (MGB) Institutional Review Board. The MGB Cutaneous Oncology Data Repository (CODR) was queried to identify patients treated within the Mass General Brigham Cancer Institute health care system between January 1, 2016, and December 31, 2025. Patients were eligible if they had a history of solid organ transplantation, a diagnosis of locally advanced or metastatic CSCC and received at least one dose of systemic ICI therapy for CSCC. The first systemic ICI administration defined the index date. Clinicopathologic, demographic, oncologic, transplant, treatment, and outcome data were abstracted from the electronic health record and de-identified using the Safe Harbor method. For multiple transplants, time to ICI was calculated from the most recent functioning allograft. Maintenance immunosuppression before ICI and after peri-ICI modification was categorized as calcineurin inhibitor, mTOR inhibitor, antimetabolite, corticosteroid, or other. Immunosuppressive modifications were classified as conversion to an mTOR-based regimen, calcineurin inhibitor reduction or discontinuation, antimetabolite reduction or discontinuation, corticosteroid intensification or pulsing, or other modification. Categories were not mutually exclusive. Incident allograft analyses excluded patients with pre-ICI rejection or graft failure. Dialysis and kidney allograft failure analyses included only kidney recipients with a functioning allograft at ICI initiation.

### Oncologic outcomes

Tumor response and best overall response were assessed according to Response Evaluation Criteria in Solid Tumors version 1.1 (RECISTv1.1). Progression-free survival (PFS) was defined as the interval from ICI initiation to disease progression or death from any cause, whichever occurred first. Overall survival (OS) was defined as the interval from ICI initiation to death from any cause. Disease-specific survival (DSS) was defined as the interval from ICI initiation to death attributable to CSCC. Deaths from other causes were censored at the date of death. Patients without an event were censored at the date of last clinical follow-up.

### Immune-related Adverse Events and Allograft Outcomes

Immune-related adverse events (irAEs) were identified through review of clinical documentation and graded according to the Common Terminology Criteria for Adverse Events, version 5.0. Events were categorized by affected organ system. Allograft rejection after ICI initiation was defined as a new clinically diagnosed or biopsy-confirmed episode of rejection occurring after the first systemic ICI administration and defined as biopsy-confirmed rejection or acute kidney injury with at least a doubling of baseline serum creatinine that was clinically attributed to rejection in the absence of an alternative explanation, as previously described.^11^ For kidney transplant recipients, graft failure was defined as irreversible loss of kidney allograft function requiring chronic dialysis.

### Statistical analysis

Baseline characteristics were summarized using descriptive statistics, including counts and percentages for categorical variables and medians with interquartile ranges for continuous variables. The Kaplan-Meier method estimated PFS, DSS, and OS. Survival probabilities and corresponding 95% confidence intervals were estimated at the 3 year prespecified landmark. The cumulative incidence of kidney allograft failure was estimated among kidney transplant recipients with a functioning allograft at ICI initiation, with death before graft failure treated as a competing event. Patients alive with a functioning allograft at last follow-up were censored at that date. Given the limited number of allograft rejection events, analyses of peri-ICI immunosuppression strategies were primarily descriptive and exploratory. Statistical analyses were performed using R version 4.6.1 (R Foundation for Statistical Computing, Vienna, Austria).

## Results

The final analytical cohort included 36 SOTRs with advanced CSCC treated with ICI. Median age at ICI initiation was 65 years (IQR, 59-69 years), twenty-nine patients (80.6%) were male, and thirty (83.3%) had an Eastern Cooperative Oncology Group Performance Status of ≤1. Thirty-four patients (94.4%) were kidney transplant recipients, with one liver and one lung transplant recipient. Median time from transplantation to ICI initiation was 9.8 years (IQR, 7.1-14.9 years). Among kidney transplant recipients, median baseline eGFR was 53 mL/min/1.73m^2^ (IQR, 34.3-70.5), and median serum creatinine was 1.3 mg/dL (IQR, 1.1-2.0) (Table 1). Maintenance immunosuppression at baseline mostly included corticosteroids in 26 patients (72.2%) and calcineurin inhibitors in 24 (66.7%). Thirteen patients (36.1%) received an mTOR inhibitor and thirteen (36.1%) an antimetabolite. Immunosuppression was modified before or at ICI initiation in 29 patients (80.6%). Modifications included corticosteroid intensification or pulsing in 20 (55.6%), calcineurin inhibitor reduction or discontinuation in 19 (52.8%), conversion to an mTOR-based regimen in 14 (38.9%), and antimetabolite reduction or discontinuation in 10 (27.8%) patients. (Table 1).

**Table 1.** Patient demographics.

| Characteristic | Total Cohort (N = 36) |
| --- | --- |
| **Demographics** |  |
| Age at ICI initiation, median (IQR), y | 65 (59-69) |
| Male sex, n (%) | 29 (80.6%) |
| ECOG PS, n (%) |  |
| 0 | 7 (19.4%) |
| 1 | 23 (63.9%) |
| 2 | 6 (16.7%) |
| Race, n (%) |  |
| White | 36 (100%) |
| Ethnicity, n (%) |  |
| Not Hispanic or Latino | 35 (97.2%) |
| **Transplant characteristics** |  |
| Transplanted organ, n (%) |  |
| Kidney | 34 (94.4%) |
| Liver | 1 (2.8%) |
| Lung | 1 (2.8%) |
| Donor Type |  |
| Deceased Donor | 11 (30.6%) |
| Living Unrelated Donor | 13 (36.1%) |
| Living Related Donor | 12 (33.3%) |
| Baseline renal allograft function, median (IQR) |  |
| eGFR among kidney transplant recipients, median (IQR), mL/min /1.73 m<sup>2</sup> | 53 (34.3-70.5) |
| Creatinine among kidney transplant recipients, median (IQR), mg/dL | 1.3 (1.1-2.0) |
| Time from transplant to ICI, median (IQR), y | 9.8 (7.1-14.9) |
| Prior allograft rejection before ICI, n (%) | 2 (5.6%) |
| **Baseline immunosuppression, n (%)** |  |
| Corticosteroid | 26 (72.2%) |
| Calcineurin inhibitor | 24 (66.7%) |
| mTOR inhibitor | 13 (36.1%) |
| Antimetabolite | 13 (36.1%) |
| Belatacept | 1 (2.7%) |
| **Immunosuppression modification before/at ICI initiation, n (%)** | 29 (80.6%) |
| Conversion to an mTOR-based regimen | 14 (38.9%) |
| Calcineurin inhibitor reduced/discontinued | 19 (52.8%) |
| Antimetabolite reduced/discontinued | 10 (27.8%) |
| Corticosteroid increased/pulsed | 20 (55.6%) |
| Other maintenance immunosuppression modified | 4 (11.1%) |
| **CSCC and treatment characteristics** |  |
| Primary tumor site, n (%) |  |
| Head and neck | 29 (80.6%) |
| Trunk | 1 ( 2.8%) |
| Extremities | 6 (16.7%) |
| AJCC 8<sup>th</sup> edition Stage Group |  |
| T - Tumor Stage, T4 | 10 (27.8%) |
| N - Nodal Stage, N2-N3 | 19 (52.8%) |
| M - Metastatic, M1 | 20 (55.6%) |
| Disease extent at ICI initiation, n (%) |  |
| Locally advanced/unresectable | 13 (36.1%) |
| Regional nodal metastases | 21 (58.3%) |
| Distant metastases | 16 (44.4%) |
| In-transit metastases, skin | 11 (30.6%) |
| Prior Therapy, n (%) |  |
| Surgery | 35 (97.2%) |
| Radiation | 28 (77.8%) |
| Chemotherapy | 10 (27.8%) |
| Cetuximab | 10 (27.8%) |
| ICI agent, n (%) |  |
| Cemiplimab | 29 (80.6%) |
| Pembrolizumab | 7 (19.4%) |

Abbreviations: AJCC: American Joint Committee on Cancer; CSCC: Cutaneous Squamous Cell Carcinoma; d: days; eGFR: estimated glomerular filtration rate; ECOG PS: Eastern Cooperative Oncology Group Performance Status; ICI: Immune Checkpoint Inhibitor; IQR: Interquartile Range; irAE: immune-related Adverse Event; mTOR: mechanistic/mammalian target of rapamycin; n: number of patients

The primary CSCC arose in the head and neck in 29 patients (80.6%). At ICI initiation, 13 patients (36.1%) had locally advanced or unresectable disease, 21 (58.3%) had regional nodal metastases, 16 (44.4%) had distant metastases, and 11 (30.6%) had in-transit cutaneous metastases. Cemiplimab was administered to 29 patients (80.6%) and pembrolizumab to 7 (19.4%) (Table 1).

Best overall response according to RECISTv1.1 and the corresponding percentage change in target-lesion burden are shown in Figure 1. At a median follow-up of 42.1 months, 9 patients (25.0%) achieved a complete response (CR), 7 (19.4%) a partial response (PR), 4 (11.1%) stable disease (SD), and 16 (44.4%) progressive disease (PD). The objective response rate was 44.4% (95%CI, 27.9%-61.9%), and the disease control rate was 55.6% (95%CI, 38.1%-72.1%). At 3 years, PFS was 18.6% (95%CI, 8.9%-38.8%), DSS was 65.8% (95%CI, 50.1%-86.4%), and OS was 32.5% (95%CI, 18.9%-55.9%) (Figure 2).

**Figure 1.**
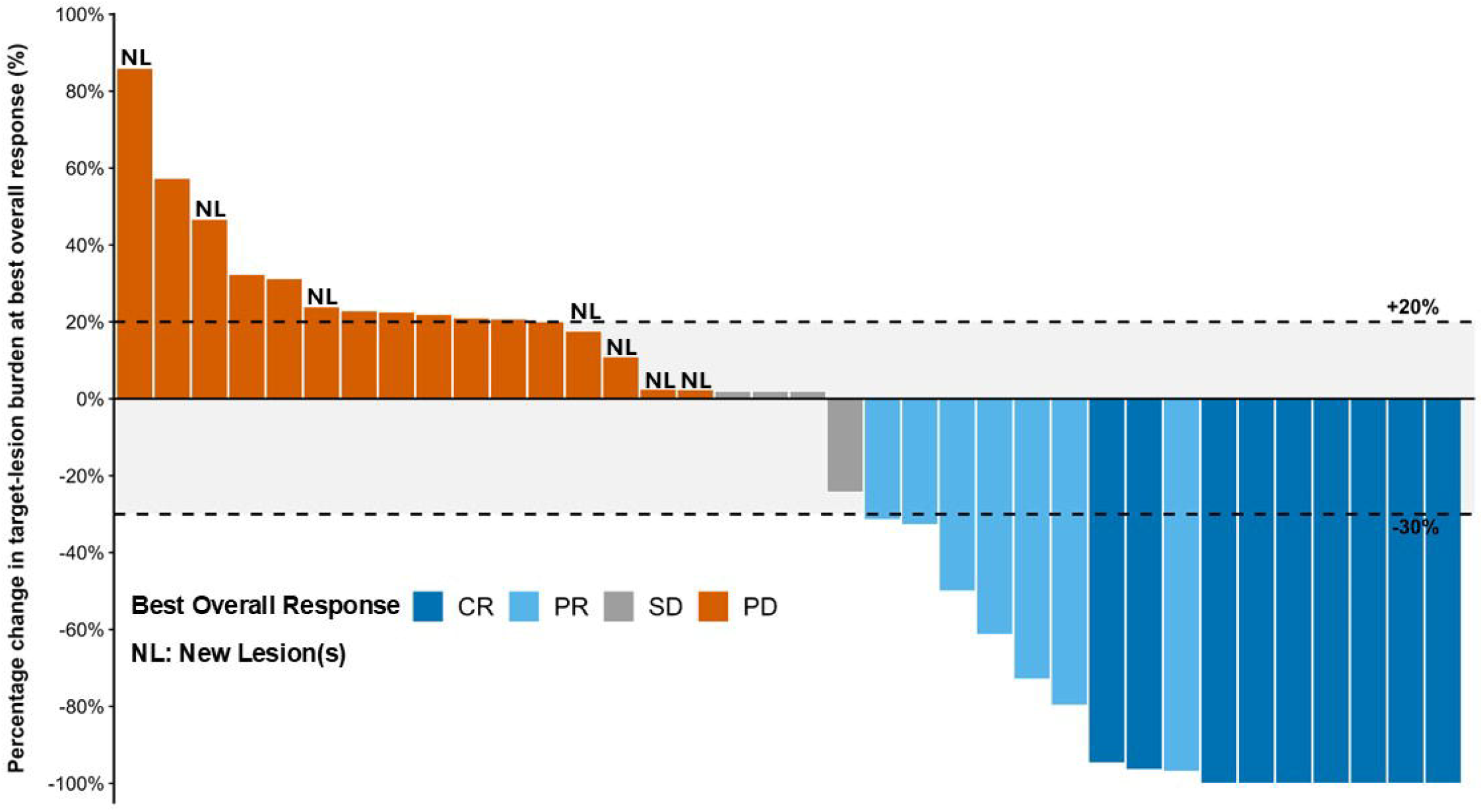
Best overall response (per RECISTv1.1) to immune checkpoint inhibition in SOTRs with CSCC. Waterfall plot showing the percentage change in target-lesion burden from baseline at the assessment corresponding to best overall response according to RECISTv1.1. Bars are colored by best overall response: complete response (CR), partial response (PR), stable disease (SD), and progressive disease (PD). Patients with PD based on a new lesion are indicated (NL). Dashed horizontal lines indicate the RECISTv1.1 thresholds for partial response (−30%) and progressive disease (+20%) based on target-lesion measurements. No cases of pseudoprogression were observed. Abbreviations: RECISTv1.1: Response Evaluation Criteria in Solid Tumors, version 1.1; CR: Complete Response; PR: Partial Response; SD: Stable Disease; PD: Progressive Disease, NL: New Lesion(s).

**Figure 2.**
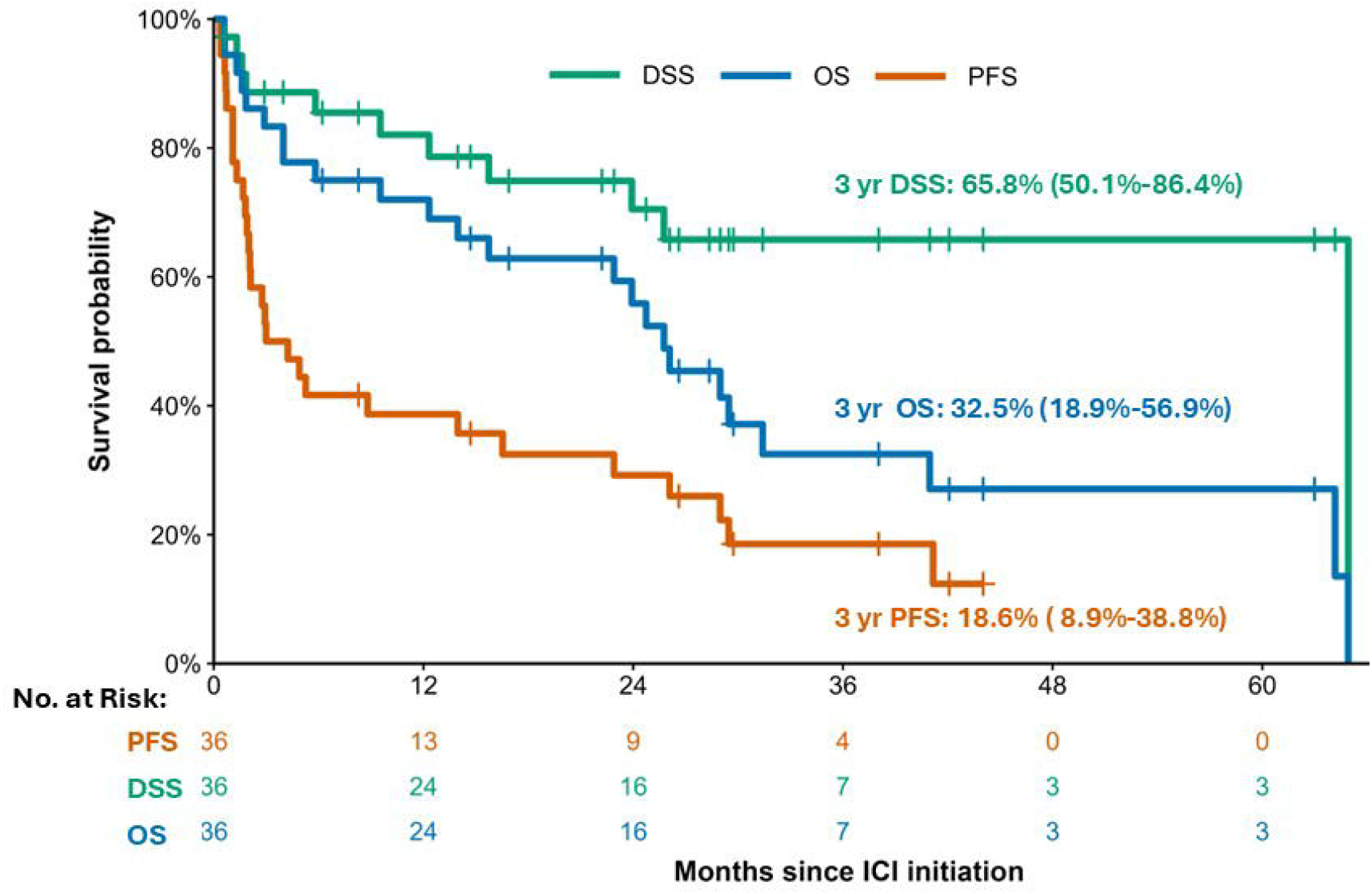
Survival outcomes in SOTRs following immune checkpoint inhibitor treatment. Kaplan-Meier estimates of progression-free survival (PFS), disease-specific survival (DSS), and overall survival (OS) were calculated from the date of first immune checkpoint inhibitor (ICI) administration. PFS was defined as time to disease progression or death from any cause, OS as time to death from any cause, and DSS as time to death attributable to cutaneous squamous cell carcinoma; deaths from other causes were censored at the date of death for DSS. Tick marks indicate censored observations, and numbers at risk are shown below the x-axis. At 3 years, PFS was 18.6% (95% CI, 8.9%-38.8%), DSS was 65.8% (95% CI, 50.1%-86.4%), and OS was 32.5% (95% CI, 18.9%-55.9%). Median follow-up was 42.1 months. Abbreviations: No.: Number; ICI: Immune Checkpoint Inhibitor; CI: Confidence Interval; DSS: Disease-Specific Survival; OS: Overall Survival; PFS: Progression-Free Survival.

At least one immune-related adverse event (irAE) of any grade occurred in 26 patients (72.2%). Eleven patients (30.6%) experienced a grade 3 or higher irAE. Renal irAEs were the most frequently recorded category, occurring in 15 patients (41.7%), including 6 (16.7%) with grade 3 or higher events.

Dermatologic, endocrine, and gastrointestinal irAEs occurred in 10 (27.8%), 9 (25.0%), and 7 patients (19.4%), respectively. Pulmonary and hepatic irAEs each occurred in 3 patients (8.3%). ICI was discontinued because of an adverse event in 8 patients (22.2%) (Table 2).

**Table 2.** Immune-related adverse events and allograft outcomes following ICI treatment for CSCC.

| **Immune-related adverse events (irAEs)** | **Any grade, n (%)** | **Grade ≥3, n (%)** |
| --- | --- | --- |
| Patients with at least one irAE | 26 (72.2) | 11 (30.6) |
| Dermatologic | 10 (27.8) | 2 ( 5.6) |
| Endocrine | 9 (25.0) | 1 ( 2.8) |
| Gastrointestinal | 7 (19.4) | 4 (11.1) |
| Hepatic | 3 ( 8.3) | 0 ( 0.0) |
| Pulmonary | 3 ( 8.3) | 1 ( 2.8) |
| Renal | 15 (41.7) | 6 (16.7) |
| ICI discontinued because of irAE(s) | 8 (22.2) | 5 (13.9) |
| **Allograft outcomes following immunotherapy treatment for CSCC<sup>a</sup>** |  |  |
| Time from ICI initiation to rejection, median (IQR), d | 84 (33-112) |  |
| Graft rejection | 5 (14.7) |  |
| Time from ICI initiation to graft loss, median (IQR), d | 84 (72-375) |  |
| Graft loss | 5 (14.7) |  |
| Dialysis initiation among kidney transplant recipients<sup>b</sup> | 5 (15.6) |  |

<sup>a</sup>Two patients experienced allograft rejection before initiation of ICI therapy and were excluded from the denominator for allograft outcomes analyses. <sup>b</sup>For analysis of dialysis initiation following ICI exposure, two patients with pre-ICI allograft rejection and two non-renal solid organ transplant recipients were excluded from the analysis. Abbreviations: d: days; ICI: Immune Checkpoint Inhibitor; IQR: Interquartile Range; irAE: immune-related Adverse Event; n: number of patients.

Two patients with pre-ICI allograft rejection were excluded from incident allograft analyses. Among 34 SOTRs at risk, 10 (29.4%) developed kidney allograft rejection a median of 84 days after ICI initiation (IQR, 33-112). Five of these 10 patients progressed to kidney allograft loss requiring chronic dialysis, representing 15.6% of the 32 at-risk kidney recipients. Four of the five patients with graft loss had biopsy-confirmed rejection. Best overall response among patients with graft loss was CR in 1, SD in 1, and PD in 3 (Table 2). Neither nonrenal SOTR developed rejection or graft failure. With death before graft failure treated as a competing event, the 3-year cumulative incidence of kidney allograft failure was 16.7% (95% CI, 2.9%-30.5%) (Figure 3).

**Figure 3.**
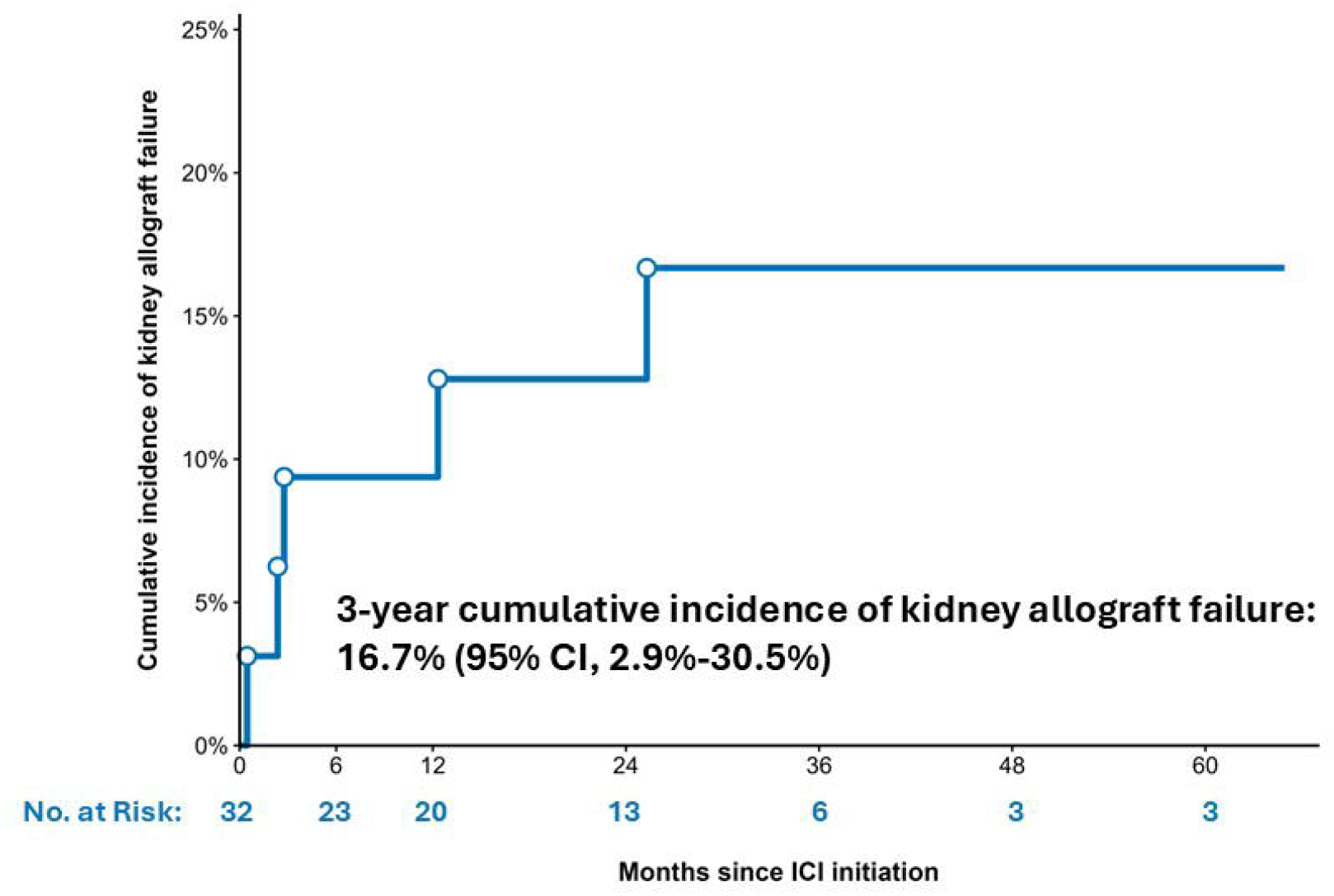
Cumulative incidence of renal allograft failure following immune checkpoint inhibitor treatment for CSCC. Cumulative incidence of renal allograft failure among kidney transplant recipients with a functioning allograft at immune checkpoint inhibitor initiation. Death before graft failure was treated as a competing event, and patients alive with a functioning graft at last follow-up were censored. Open circles indicate graft-failure events. Numbers at risk are shown below the x-axis in blue. The 3-year cumulative incidence of kidney allograft failure was 16.7% (95% CI, 2.9%-30.5%). Abbreviations: No.: Number; ICI: Immune Checkpoint Inhibitor; CI: Confidence Interval.

## Discussion

The role of systemic ICI treatment in SOTRs with advanced CSCC remains uncertain, with evidence drawn largely from case reports, small series, and prospective studies. Our data represents one of the largest cohorts reported to date. We identified 36 SOTRs at our institution, over a 9 year period, who were treated with systemic ICI for advanced or metastatic CSCC. At a median follow-up of 42.1 months, the objective response rate was 44.4%, including a complete response in 25% of the patients. In addition, we observed a disease control rate of 55.6%. This compares favorably to the response rates observed with ICI in non-transplant patients with advanced CSCC where objective responses have generally been around 47-50%.^9^ Acute rejection occurred in 10 (29.4%) of the patients with a 16.7% 3-year cumulative incidence of renal allograft failure. Thus, in a real-world population of SOTRs, systemic ICI appeared to provide clinically meaningful cancer control and was associated with a measurable but nonuniversal risk of irreversible graft failure. Our study provides a longitudinal assessment of oncologic and allograft loss outcomes beyond the relatively short observation periods previously reported.^12–14^

Longitudinal CSCC-specific data on graft loss remain limited. Murakami et al. reported acute rejection in 42% of 69 ICI-treated transplant recipients with various cancers; 65.5% of those with rejection progressed to graft failure. Although 9/24 (37.5%) patients with CSCC developed rejection, subgroup graft-loss outcomes were not reported.^11^ An individual-participant meta-analysis across cancer types estimated a 1-year graft-loss incidence of 18.4%, and greater rejection risk with melanoma than CSCC (HR, 2.88; 95% CI, 1.69-4.90).^15^ Differences in cancer type, outcome definitions, and study design limit direct comparisons. We defined kidney graft failure as irreversible loss of allograft function requiring chronic dialysis. This definition distinguishes permanent organ loss from creatinine elevation, acute kidney injury, or rejection that responds to treatment. All five graft losses followed rejection, and four of these five patients had biopsy-confirmed rejection, reinforcing the distinction between rejection episodes and irreversible graft failure. Competing-risk analysis provided a more appropriate long-term graft-loss estimate than a crude proportion. These distinctions are essential when counseling patients about rejection and the risk of permanent organ loss and coordinating individualized multidisciplinary care.

In nontransplant patients, irAEs commonly involve the skin, lungs, liver, gastrointestinal tract, and endocrine organs, whereas renal involvement is less frequent. Renal irAEs have been reported in 1.4%-4.9% of patients receiving anti-PD-1/PD-L1 monotherapy outside transplantation^.16^ In our cohort, renal irAEs were the most frequent, affecting 15 patients (41.7%). The high renal irAE rate may reflect our predominantly renal cohort or increased allograft susceptibility. We likely treated more kidney recipients because of higher CSCC incidence and dialysis availability if graft failure occurred. Renal irAEs may present as AKI, mimicking rejection or transient renal dysfunction. Although both may require immunosuppression, early rejection evaluation and renal biopsy, when appropriate, are important to distinguish these processes and guide management.

Three-year PFS, DSS, and OS were 18.6%, 65.8%, and 32.5%, respectively. The divergence between DSS and OS suggests that cancer control is only one determinant of survival in SOTRs, who remain vulnerable to cardiovascular disease, infection, impaired allograft function, and complications of chronic immunosuppression^17,18,19^ Despite meaningful initial responses, low 3-year PFS highlights the limited proportion remaining alive without progression. Further studies should characterize response durability and patterns of progression in SOTRs. Favorable DSS should not obscure the substantial medical burden that persists after cancer control. Disease-specific outcomes alongside OS may provide a more complete assessment of treatment benefit.^18–21^

Optimal maintenance immunosuppression during ICI treatment remains unresolved. Most patients underwent individualized peri-ICI modifications, including transition to mTOR inhibitors, calcineurin inhibitor reduction or discontinuation, antimetabolite reduction, and corticosteroid intensification or pulsing. These strategies overlapped and were not protocolized. In a phase I trial, 12 kidney recipients with advanced CSCC received cemiplimab after mTOR conversion, with pulsed prednisone followed by maintenance dosing. No rejection or graft loss occurred over a median follow-up of 6.8 months, although one death was attributed to the mTOR inhibitor.^12^ We did not routinely use pulsed corticosteroids during ICI treatment. Heterogeneous immunosuppression and only five graft-loss events preclude conclusions about regimen superiority. Prospective studies with standardized regimens and longer follow-up are needed to determine how immunosuppression modification influences antitumor activity and allograft outcomes.

Limitations include retrospective, single-institution design and potential selection and information bias. The modest cohort and few allograft events precluded robust comparisons of immunosuppressive regimens or multivariable modeling of rejection risk. Attribution to ICI remains uncertain, particularly for later events, because clinical or histopathologic confirmation establishes rejection without establishing its cause. All patients received anti-PD-1 agents. The favorable immune-related safety profile reported with cosibelimab in CSCC warrants evaluation in SOTRs, although comparative toxicity and allograft rejection risk remain undefined.^22^ Kidney recipients predominated, potentially reflecting our greater willingness to consider ICI when dialysis was available after graft failure. Dialysis provides replacement of kidney function but does not reverse allograft loss. Outcomes therefore may not generalize to other organ recipients. Nevertheless, the CSCC-specific cohort, extended follow-up, and distinction between rejection and irreversible graft loss provide complementary evidence to prior heterogeneous series and small prospective studies.[

For selected patients with sustained oncologic remission after kidney graft failure, reassessment of transplant candidacy may eventually be appropriate. The optimal disease-free interval, immunosuppressive strategy for a subsequent graft, and risks of recurrent malignancy or renewed alloimmune injury after checkpoint blockade remain unknown. As prospective follow-up accumulates, these questions merit discussion between oncology and transplant teams, particularly for patients with durable cancer control who remain dependent on dialysis.

In conclusion, ICI may provide meaningful cancer control in SOTRs with advanced CSCC, but treatment carries a clinically relevant risk of rejection and irreversible graft failure. Further studies across organ types are needed to define treatment outcomes and optimize immunosuppression. Our findings support individualized, multidisciplinary care that integrates antitumor efficacy, allograft preservation, and long-term survivorship.

## Data Availability

All aggregate data supporting the findings of this study are contained within the manuscript. Individual-level patient data are not publicly available because of patient privacy and institutional restrictions but may be available from the corresponding author upon reasonable request and with necessary institutional approval.

